# Vaccine-induced human monoclonal antibodies to PfRH5 show broadly neutralizing activity against *P. falciparum* clinical isolates

**DOI:** 10.1101/2024.05.24.24305116

**Authors:** Laty G. Thiam, Kirsty McHugh, Aboubacar Ba, Rebecca Li, Yicheng Guo, Mariama N. Pouye, Awa Cisse, Dimitra Pipini, Fatoumata Diallo, Seynabou D. Sene, Saurabh D. Patel, Alassane Thiam, Bacary D. Sadio, Alassane Mbengue, Inés Vigan-Womas, Zizhang Sheng, Lawrence Shapiro, Simon J. Draper, Amy K. Bei

**Affiliations:** G4 - Malaria Experimental Genetic Approaches & Vaccines, Pôle Immunophysiopathologie et Maladies Infectieuses, Institut Pasteur de Dakar, Dakar, Senegal.; Department of Biochemistry and Kavli Institute for Nanoscience Discovery, University of Oxford, Oxford, UK.; Department of Epidemiology of Microbial Diseases, Yale School of Public Health, New Haven, CT, USA.; Aaron Diamond AIDS Research Center, Columbia University Vagelos College of Physicians and Surgeons, New York, NY, USA.; Pôle Virologie, Institut Pasteur de Dakar, Dakar, Senegal.; Department of Biochemistry and Biophysics, Columbia University, New York, NY, USA.; NIHR Oxford Biomedical Research Centre, Oxford, UK.

## Abstract

Vaccines to the *Plasmodium falciparum* reticulocyte binding-like protein homologue 5 (PfRH5) target the blood-stage of the parasite life cycle. PfRH5 has the potential to trigger the production of strain-transcendent antibodies and has proven its efficacy both in pre-clinical and early clinical studies. Vaccine-induced monoclonal antibodies (mAbs) to PfRH5 showed promising outcomes against cultured *P. falciparum* laboratory strains from distinct geographic areas. Here, we assessed the functional impact of vaccine-induced anti-PfRH5 mAbs on more genetically diverse *P. falciparum* clinical isolates. We used mAbs previously isolated from single B cells of UK adult PfRH5 vaccinees and used ex-vivo growth inhibition activity (GIA) assays to assess their efficacy against *P. falciparum* clinical isolates. Next-generation sequencing (NGS) was used to assess the breadth of genetic diversity in *P. falciparum* clinical isolates and to infer the genotype/phenotype relationship involved in antibody susceptibility. We showed a dose-dependent inhibition of clinical isolates with three main GIA groups: high, medium and low. Except for one isolate, our data show no significant differences in the mAb GIA profile between *P. falciparum* clinical isolates and the 3D7 reference strain, which harbors the vaccine allele. We also observed an additive relationship for mAb combinations, whereby the combination of GIA-low and GIA-medium antibodies resulted in increased GIA, having important implications for the contribution of specific clones within polyclonal IgG responses. While our NGS analysis showed the occurrence of novel mutations in the *pfrh5* gene, these mutations were predicted to have little or no functional impact on the antigen structure or recognition by known mAbs. Our present findings complement earlier reports on the strain transcendent potential of anti-PfRH5 mAbs and constitute, to our knowledge, the first report on the susceptibility of *P. falciparum* clinical isolates from natural infections to vaccine-induced human mAbs to PfRH5.

## Introduction

Malaria continues to afflict humankind, with over 200 million cases and more than 600,000 deaths reported in 2022^1^. Combined efforts in both preventive and therapeutic measures have helped reduce the malaria burden to more than a third over the last two decades^2,3^. This progress has however stalled since 2015 with an observed plateau^1^. Moreover, recent data showed an increase of 5 million cases globally between 2021 and 2022^1^. With the growing threats associated with the emergence and spread of parasite and vector resistance to antimalarials and insecticides, respectively^4^, this situation will likely worsen if new strategies are not developed soon. While a very high efficacy malaria vaccine would be a key transformative tool in the malaria elimination toolbox, achieving this goal has been challenged by both the complexity of the parasite life cycle and the extent of genetic diversity, which can limit vaccine efficacy^5^.

Infection with *Plasmodium falciparum*, the most lethal species of human malaria parasites, starts with sporozoites inoculated through the bite of an infected female *Anopheles* mosquito. This stage is targeted by RTS,S/AS01 (Mosquirix™)^6^ and R21/Matrix-M™^7^, so far, the only two malaria vaccines recommended by the World Health Organization (WHO) for use in children living in areas with moderate to high malaria transmission^1^. In phase 3 efficacy trials, the protective efficacy against clinical malaria of R21/Matrix-M, over a 12-month follow up period, was 67-75% in children (aged 5-36 months) who were fully vaccinated (3 doses, efficacy endpoint measured at 12 months after the third dose)^7^. On the other hand, phase 3 clinical assessment of RTS,S/AS01 protective efficacy against uncomplicated, over a median 48-months follow up, was respectively 18.3% and 28.2%in infants (6-12 weeks) and children (5-17 months)^6^. Moreover, in both age groups a booster dose at 20 months after the third dose yielded efficacy against uncomplicated clinical malaria of 25.9% in infants and 36.3% in children^6^. A highly effective malaria vaccine that provides durable protection is urgently needed to eliminate malaria. Consequently, a strain-transcendent blood-stage vaccine could complement the pre-erythrocytic efficacy achieved through RTS,S/AS01 and/or R21/Matrix-M™ vaccination. Blood-stage vaccines have been explored in the past, but have often struggled to induce strain-transcendent strongly efficacious immunity. AMA1-based vaccines induce strong antibody responses, however, fail to provide significant protection against clinical malaria and are associated with a limited efficacy in both early clinical trials^8–10^ and control human malaria infection studies^11^. AMA1 vaccination has been shown to provide a modest efficacy against clinical malaria (17.4%). This lack of efficacy is attributed to the strain-specific immunity triggered upon AMA1 vaccination, which selects for breakthrough infections from non-matched genotypes circulating in the natural parasite population, as efficacy against clinical malaria with vaccine-type AMA1 was 64.3%^12^. Likewise, this allele-specific immunity has also been associated to the partial efficacy of RTS,S following phase 3 clinical trial studies^13^. This highlights the potential impact of current efforts in the development of next-generation blood stage vaccines, such as the *P. falciparum* reticulocyte binding protein homologue 5 (PfRH5) that have shown promise in eliciting strain-transcending immune responses^14^. Early testing of the effect of naturally arising *P. falciparum* genetic diversity on PfRH5 antibody responses is therefore critical to help achieve this goal.

Vaccines targeting the blood-stage of the parasite life cycle are of particular interest as they target the clinical stage of infection and are aimed at reducing parasite density and severe disease. The PfRH5 protein has emerged as a leading vaccine candidate, with attractive characteristics being both an essential and conserved^15,16^ promising blood-stage vaccine candidate. PfRH5 is by far the most advanced targeted blood-stage vaccine candidate that has reached clinical development over the last few years^17,18^. In addition to being strain-transcendent^14,19–21^, antibodies to PfRH5 have shown protection in non-human primates against heterologous blood-stage challenge^22,23^. Clinically, PfRH5 immunization triggers higher antibody titers in vaccinated infants and children relative to adults^18^. Notably, antibodies to PfRH5 are associated with a delayed parasite onset in a controlled human malaria infection (CHMI) in naïve adults, which also correlates with in vitro growth inhibition activity (GIA) of vaccine-induced antibodies^24^. Moreover, human monoclonal antibodies (mAbs) isolated from PfRH5-vaccinated UK adults have been reported with broadly neutralizing activities against *P. falciparum* laboratory lines^25^, however, their potential has not been tested on more diverse clinical parasite isolates circulating in endemic settings.

Here, we assess the inhibitory potential of vaccine-induced anti-PfRH5 human mAbs against genetically diverse *P. falciparum* clinical isolates from a malaria-endemic setting in Senegal. Parasite isolate susceptibility to antibody inhibition was performed using a flow cytometry-based *ex-vivo* GIA assay and the breadth of *PfRH5* genetic diversity was assessed using targeted deep amplicon sequencing of *PfRH5* amplicons. The potential impact of PfRH5-associated genetic diversity was predicted through structural threading of the identified polymorphisms onto the three-dimensional structure of PfRH5 in complex with its binding partner PfCyRPA, its erythrocyte receptor, Basigin, or the human mAbs used in the study.

## Results

### *P. falciparum* clinical isolates show broad susceptibility to PfRH5 vaccine-induced mAbs

We used a panel of previously characterized human mAbs isolated from single-cell-sorted plasmablasts of UK adult volunteers immunized with a viral-vectored PfRH5-based vaccine^25^. Additionally, we included two well-characterized anti-PfRH5 mouse mAbs chimeric with human Fc (c2AC7 and c9AD4)^22,26^. To assess the susceptibility of *P. falciparum* clinical isolates to PfRH5-vaccine-induced antibodies, we tested four different concentrations (150, 50, 25 and 10 µg/ml) of the mAbs on freshly collected *P. falciparum* clinical isolates from malaria symptomatic patients using a flow cytometry-based *ex-vivo* GIA assay^27^. These concentrations were chosen to incorporate the EC_50_ values of the most potent mAbs that were previously tested against laboratory lines^25^.

*P. falciparum* clinical isolates were co-incubated with mAbs during the first *ex-vivo* cycle and seeded at parasitemia of 0.5-1%. Cultures were harvested upon complete reinvasion of the parasites and the data acquired by flow cytometry. At these concentrations, the clinical isolates showed a dose-dependent susceptibility to the individual mAbs, with the chimeric mAbs, c2AC7 and c9AD4, showing the highest inhibition rates across isolates resulting in mean percent GIAs of 80.9 and 77.6% and 53.9 and 50.5%, respectively at the highest and lowest concentrations. The human vaccine-induced mAbs, R5.004, R5.016 and R5.017 showed the highest inhibition across all concentrations, with the least inhibitory mAbs being R5.001 and R5.007, while R5.008 and R5.011 showed intermediate phenotypes (**Fig. 1**); this is in line with previously reported data using the 3D7 reference laboratory line^25^. Based on these results, we stratified the mAbs into three groups referred to as GIA-high (GIA ≥ 75% at 150 µg/ml), GIA-medium (75% > GIA ≥ 50% at 150 µg/ml) or GIA-low (GIA < 50% at 150 µg/ml). Notably, the trend of the parasites’ susceptibility to individual mAbs was conserved across all test concentrations, with a relatively similar inhibition profile observed across all isolates. The only exception to this trend was isolate 400113, which showed a reduced percent GIA across all mAbs and test concentrations (**Fig. 2**).

**Figure 1:**
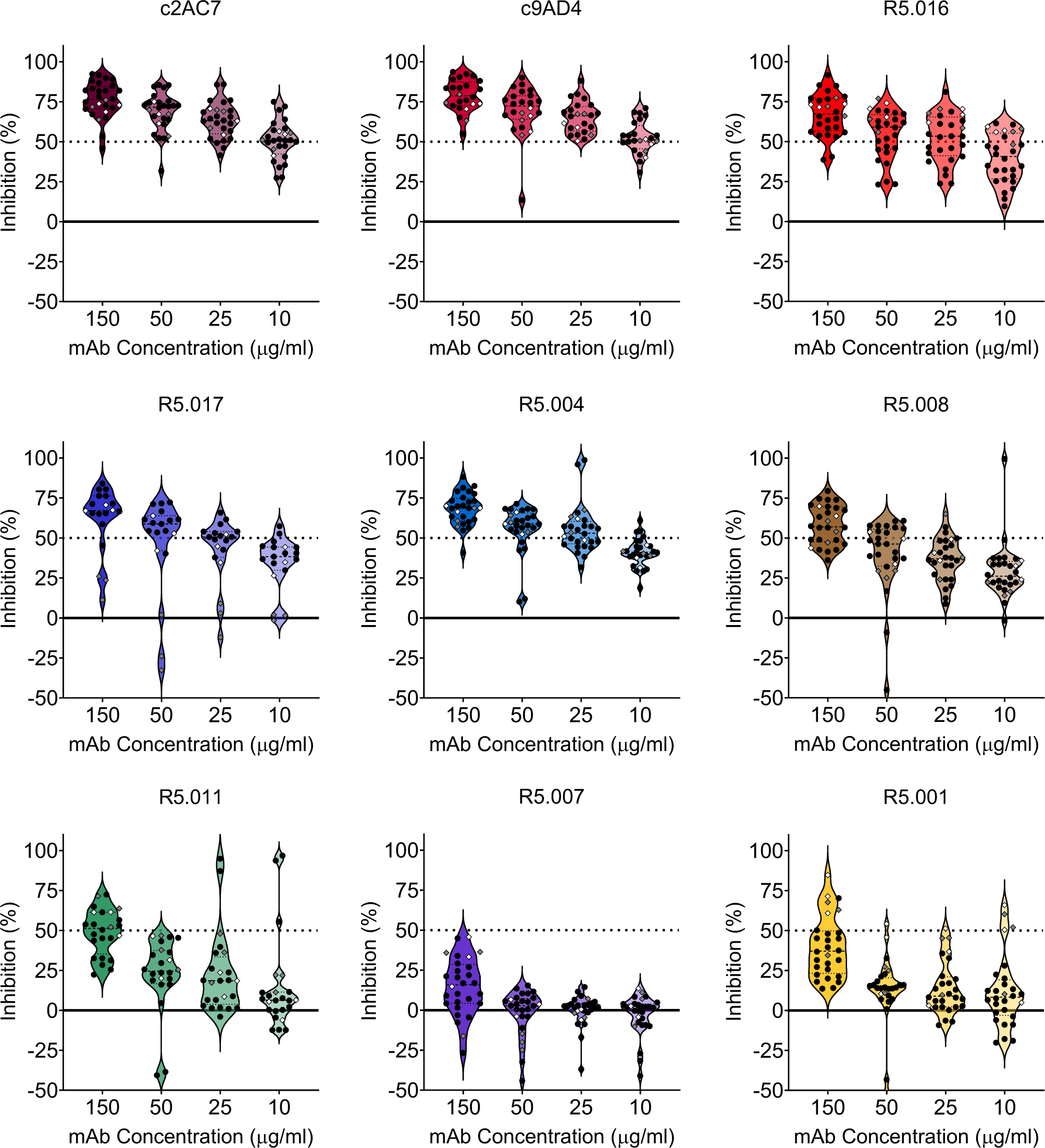
*Ex-vivo* assessment of *P. falciparum* clinical isolates’ susceptibility to anti-PfRH5 mAbs. Growth inhibitory activities (GIA) of the panel of both vaccine-induced human and chimeric mouse mAbs were tested at different concentrations against *ex-vivo* cultured *P. falciparum* clinical isolates (black dots) or *in-vitro* cultured laboratory lines (diamond shapes). Shown within the violin plots are the mean percent GIA (dashed lines) and the 25^th^ and 75^th^ quartiles (dotted lines) of the clinical isolates. Colors were chosen to specifically match that of the previously reported epitope bins of the respective mAbs^25^.

**Figure 2:**
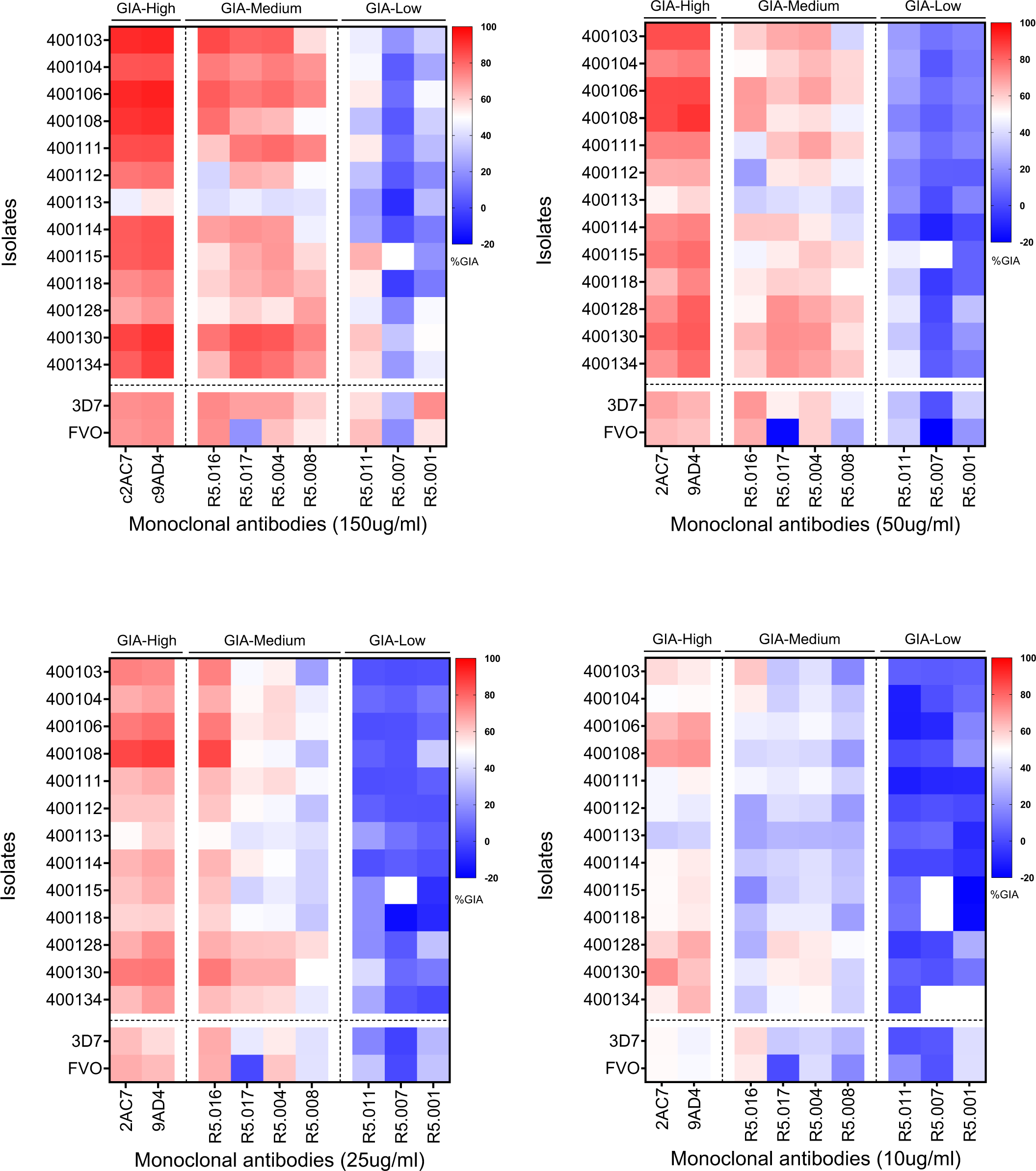
Classification of the GIA profiles of PfRH5-vaccine-induced mAbs against *P. falciparum* clinical isolates. Heat maps presenting the percent GIA data of mAbs to PfRH5 tested at different concentrations (150; 50; 25 and 10 µg/ml). The GIA data from Figure 1 were used to classify the mAbs in this panel into three main groups, GIA-high (GIA ≥ 75% at 150 µg/ml), GIA-medium (75% > GIA ≥ 50% at 150 µg/ml) or GIA-low (GIA < 50% at 150 µg/ml) separated by vertical dotted lines. Horizontal dotted lines depict the separation between *P. falciparum* clinical isolates and laboratory lines, presented in the y-axis of the graphs.

To further ascertain the clinical isolates’ susceptibilities to the panel of mAbs, we compared their percent GIA data with that of the laboratory lines 3D7 (reference strain) and FVO, for which well characterized phenotypic data have been published^33^. GIA data for both 3D7 and FVO were acquired using the same batches and concentrations of mAbs and, for each strain, three independent assays were performed in duplicates on different days. We used two-way ANOVA analysis to assess the differences in percent GIA across isolates and within each test concentration and found no significant differences in mean percent GIA among the population of clinical isolates relative to 3D7. The two-way ANOVA was associated to the Dunnett’s multiple comparison test to assess the pairwise differences in %GIA between 3D7 and the individual clinicals isolates. This analysis revealed only two isolates (400113 and 400130) showing significant differences in %GIA of clinical isolates relative to 3D7 at the highest concentration (150ug/ml), while these differences were more pronounced at the lower concentrations with 13, 11 and 12 isolates showing significant %GIA relative to 3D7, respectively at 50, 25 and 10ug/ml concentrations. Of these isolates, only 400103 showed significant difference across all test concentrations. Encouragingly, none of the antibodies showed significant differences across all test concentrations, (**Supplemental Table 1**). When comparing FVO to the reference 3D7, significant differences in percent GIA were observed across all concentrations in the presence of the R5.017 mAb, thereby confirming an earlier report in which this difference was attributed to the immune escape phenotype associated with the PfRH5 S197Y mutation in the FVO strain^25^. Otherwise, significant differences in percent GIA between FVO and 3D7 were only observed when comparing the results for mAb R5.001 at 150 µg/ml (*p = 0.046*) and the R5.008 at 50 µg/ml (*p = 0.025* (**Supplemental Table 1**).

Taking into consideration the earlier observed immune evasive phenotype of FVO relative to 3D7 in the presence of the R5.017 mAb, we defined a conservative range of values for the variability inherent in the assay, using our replicate data from 3D7. We defined a GIA difference as biologically relevant (i.e. loss of GIA susceptibility) if an isolate demonstrated a percent GIA different from that of the mean percent GIA of 3D7 ± 3 SD (standard deviations) from the 3 experimental replicates for any mAb test concentration. Such an approach was previously devised to determine the error of the assay for GIA in the presence of anti-PfRH5 antibodies^28^. Of note, here we did not include many factors that could increase variability such as blood donors, etc. and as such our cut-offs may be very conservative. Encouragingly, while we observed some variation in the clinical isolates’ susceptibility to anti-PfRH5 mAbs, none of the isolates showed a complete loss of GIA susceptibility across all mAbs tested here (**Fig. 3**). Interestingly, R5.016 showed the greatest variability in GIA and multiple isolates showed lower GIA activity across all test concentrations (**Fig. 3 and Supplemental Fig. 1A-C**). Moreover, at the highest concentration (150 µg/ml), both R5.016 and R5.004 showed multiple isolates with reduced GIA susceptibilities, while the percent GIA of the bulk of these isolates (2 of 7 for R5.016 and 5 of 6 for R5.004) was at the borderline of the set threshold (**Fig. 3**).

**Figure 3:**
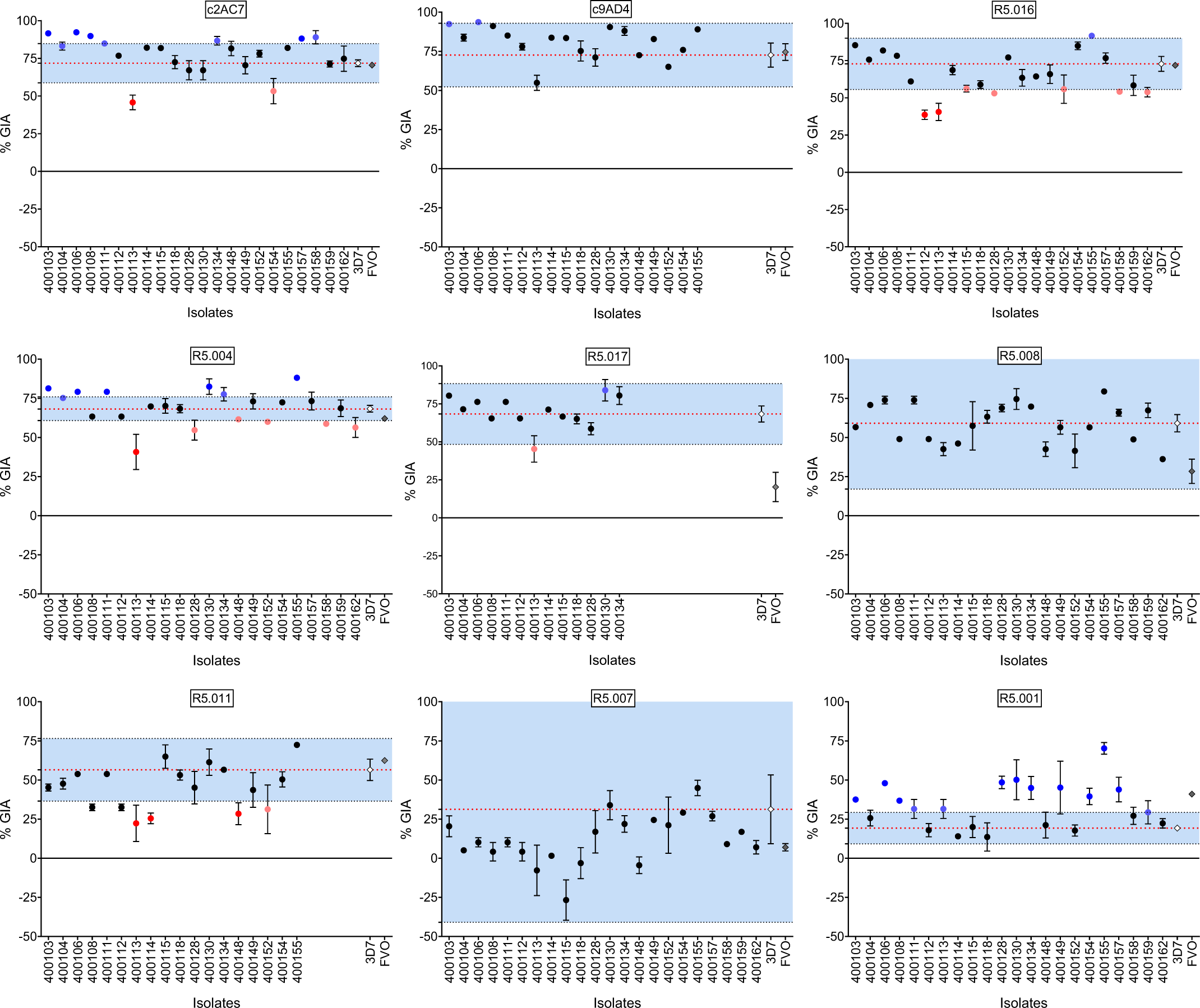
Variation of percent GIA ranges and immune susceptibility profiles of *P. falciparum* clinical isolates to PfRH5-vaccine-induced mAbs at 150µg/ml. Dot plots of %GIA to anti-PfRH5 mAbs (Y-axis) of *P. falciparum* clinical isolates (X-axis). The %GIA susceptibility ranges in blue are defined as the mean 3D7 %GIA (red dotted line) ± 3SD 3D7 %GIA (black dotted lines). Black dots represent isolates laying within the defined susceptibility ranges. Red dots represent isolates reflecting a reduced GIA susceptibility phenotype (bright red) or laying at the borderline (dim red) of the lower limit of the defined susceptibility threshold. Blue dots represent isolates showing an increased GIA susceptibility (bright blue) or laying at the borderline (dim blue) of the upper limit of the defined susceptibility threshold.

### Antibody combinations enhance GIA activity against *P. falciparum* clinical isolates

Having shown the broad neutralizing activities of PfRH5-vaccine-induced antibodies on *P. falciparum* clinical isolates, we sought to investigate the earlier reported synergistic property of R5.011 relative to mAbs with GIA-medium and GIA-low phenotypes. In these assays, R5.011 was combined in equal concentrations (150 µg/ml each) with either R5.001, R5.007 or R5.008 in *ex-vivo* GIA assays or *in vitro* GIA assays of *P. falciparum* clinical isolates or laboratory lines, respectively. Notably, there was a significant increase in the mean percent GIA of the antibody combinations relative to that of R5.001, R5.007 or R5.008 alone (*p < 0.0001*; Wilcoxon matched-pairs signed rank test). The same was true when comparing the mean percent GIA of the antibody combinations to that of R5.011 alone. Similar to the single mAb data, there were no significant differences between the mean percent GIA of clinical isolates to that of 3D7, however, there was a greater range of GIA values for the clinical isolates (**Fig. 4A**). Across all isolates, the combination R5.011-R5.007 showed more significant differences in GIA profiles relative to 3D7, while only two isolates, 400113 and 400128 showed significantly lower GIA profiles relative to 3D7 across all combinations (One-way ANOVA and Dunnett’s multiple comparison test) (**Fig. 4B**). Having shown that R5.011 was able to potentiate the activities of the least potent mAbs in inhibiting growth of *P. falciparum* clinical isolates, we next sought to investigate whether these combinations were synergistic and, given the limited number of test concentration combinations, we opted for the Bliss additivity model as described previously^29^. The predicted Bliss additivity percent GIA was calculated from the percent GIAs of the individual mAbs in a given combination and compared to the percent GIA of the corresponding test combination. Differences between predicted and actual percent GIA data were classified as additive (Difference %GIA ≥ 10) or antagonistic (Difference %GIA ≤ −10). Notably, these combinations mostly resulted in an additive interaction between the mAbs against most of the isolates tested here, except for 400128 where all combinations were antagonistic (% GIA Difference −20.98 (p = 0.002; t-test), −12.67 (p = 0.006; t-test) and −47.91 (p = 0.002; t-test), respectively for R5.011-R5.001, R5.011-R5.007 and R5.011-R5.008). Across all isolates, the combination of R5.001-R5.011 resulted in more additive interactions, while only two isolates (400108 and 400114) showed additivity in all combinations (**Fig. 4C-D**).

**Figure 4:**
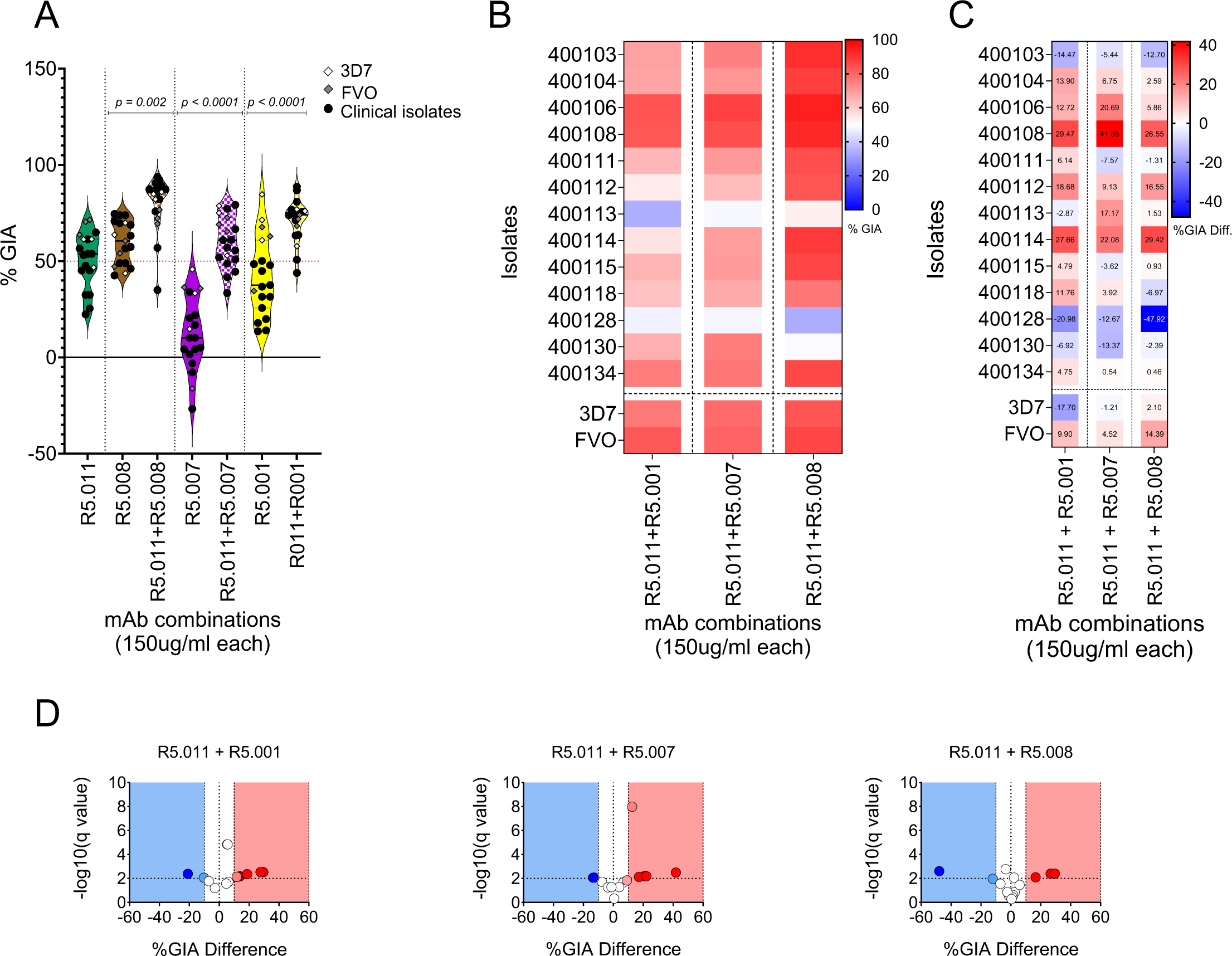
Assessment of GIA in combinatorial assays of anti-PfRH5 mAbs tested against *P. falciparum* clinical isolates. (**A**) Violin plots showing percent GIA from combinations of R5.011 (green) with either R5.001 (yellow), R5.007 (purple) or R5.008 (brown). Depicted in the plots are the mean percent GIA (plain black line) and the 25^th^ and 75^th^ quartiles (dotted black lines). Here, mAbs were combined in equal concentrations and incubated with equal volumes of either *P. falciparum* clinical isolates (black dots) or laboratory lines (diamond shapes). The resulting GIA was compared to the single antibody at the same concentration. All assays were performed in duplicates and the data are presented as means from the two replicates. Statistical differences between antibody combinations and single antibody treatments were computed in GraphPad Prism using the Wilcoxon matched-pairs signed rank test. (**B**) Heat map showing the GIA profiles of antibody combinations (x-axis) against *P. falciparum* parasites (y-axis). The dotted vertical lines separate the different antibody combinations while *P. falciparum* clinical isolates are separated from the laboratory lines by the horizontal dotted line. (**C**) heat maps and (**D**) Volcano plots depicting the difference between the Bliss additivity predicted percent GIA and the actual percent GIA from mAb combinations. Bliss analysis was calculated as previously described^35^ and the multiple unpaired t-test was used to assess the statistical difference. Volcano plots show -log10 of the q-values.

### Next-generation sequencing shows a high level of PfRH5 genetic diversity in *P. falciparum* clinical isolates from Kédougou

Given the extreme diversity of malaria parasites and its potential to impact on vaccine efficacy and immune evasion, we further assessed the breadth of PfRH5-associated polymorphisms, in addition to the MOI per isolate in our parasite population. PfRH5 genomic sequences were amplified from individual samples and subjected to next-generation sequencing using the Illumina NovaSeq 6000 platform, while the *msp1* and *msp2* genes were used to assess the number of clones per isolate. PfRH5-associated single nucleotide polymorphisms (SNPs) were called relative to the 3D7 reference using the Geneious prime software. The median MOI of our study population was 2.59, with individual patient infections harboring 1 to 5 distinct *P. falciparum* genotypes (**Fig. 5A**). Interestingly, our data showed the wild-type allele to be the minor allele, representing only 27.3% of our study population, while 72.7% of the parasites presented at least one SNP in the PfRH5 locus (**Fig. 5B**). Overall, a total of twelve SNPs was identified within our parasite population, with the C203Y being the most frequent identified in 59.1% of isolates, while the majority of the other identified SNPs were rare and identified in single isolates (4.5%) (**Fig. 5A-B**). Of these SNPs, five were novel (K51R, I60V, D243N, F505Y and Y254C) and all but one (K51R) occurred in a single isolate (**Fig. 5B**). We next sought to assess the allele frequencies within complex samples. To this end, the variant read frequencies herein reported were calculated as the frequency of the number of reads with any given SNP over the total number of reads mapped to the reference sequence at the SNP position. As seen with the SNP prevalence, the highest variant read frequencies were associated with the C203Y mutation, with frequencies ranging from 4.1 to 100%. Interestingly, of the novel SNPs, D243N presented the highest variant read frequency (99.7%) and was present in a monogenomic infection (MOI = 1) representing the least susceptible isolate 400113 (**Fig. 5C**). However, except for C203Y, none of the SNPs reported here occurred at a high prevalence and high variant frequency at the same time (**Fig. 5B-C**).

**Figure 5:**
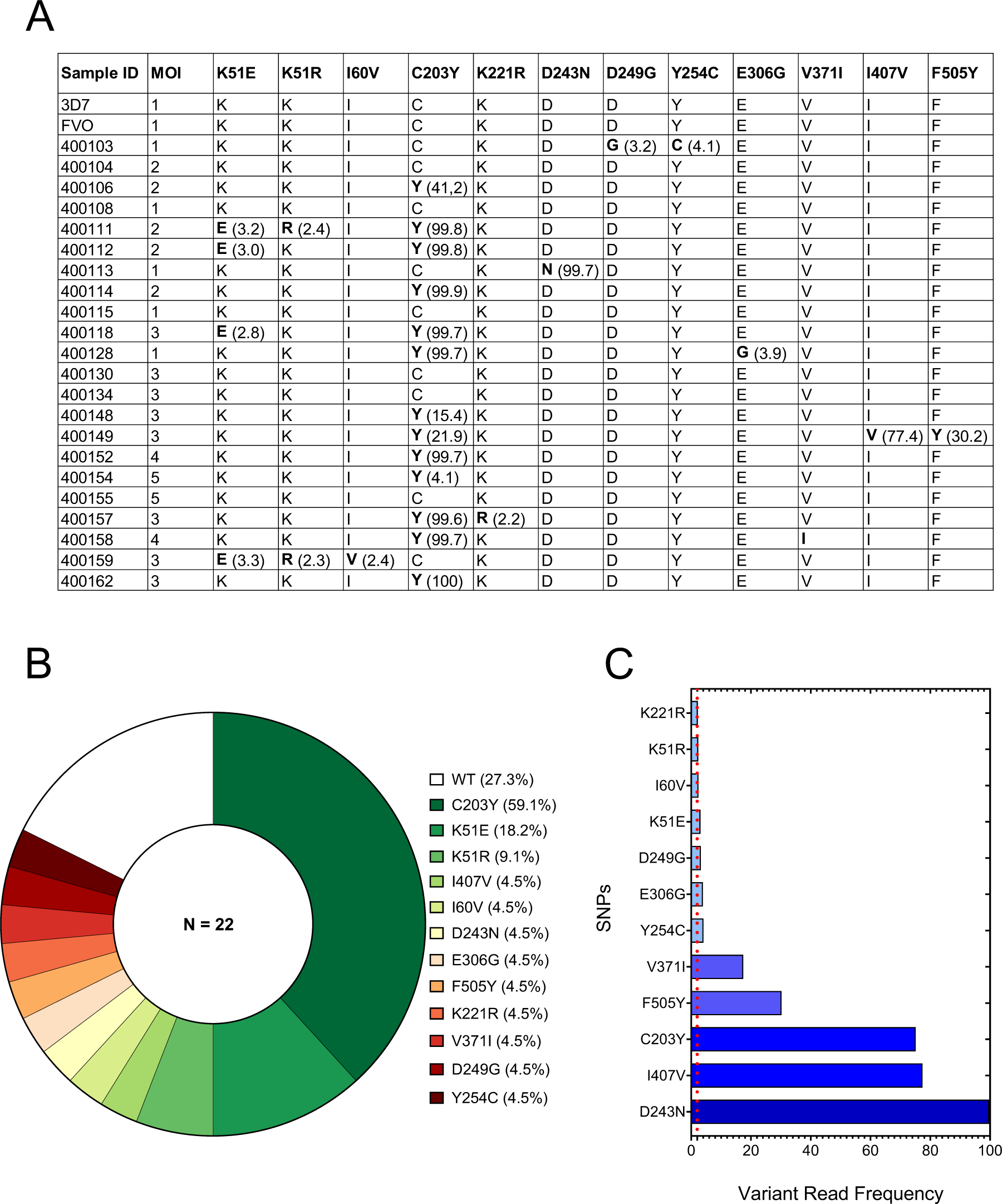
PfRh5-associated genetic diversity in *P. falciparum* clinical isolates from Kédougou. (**A**) Summary table showing the multiplicity of infections (MOI) per isolate determined through *msp1-2* typing as well as the number of SNPs at each position (bold characters) and their respective read frequencies (shown in brackets). (**B**) The prevalence of PfRH5-associated SNPs was calculated as the percentage of SNPs detected within the total sample population (N=22). PfRH5 sequencing was performed from *pfrh5* amplicons using the Illumina NovaSeq 6000 sequencing platform and variant analysis was performed using the Geneious Prime software version 23.1.1. (**C**) SNP frequency was determined from the sequencing data outputs and calculated as the percentage of the variant read coverage relative to the coverage at the variant position. The red dotted line depicts the 5% threshold frequency, arbitrarily defined here as a cut-off between low and high-frequency SNPs. The graphs were plotted using the GraphPad Prism version 1.0.2 software.

### Structure-based insights on potential SNP function

Taking into consideration the existing relationship between genetic diversity and immune evasion, as seen in the case of the PfRH5 S197Y mutation associated with the FVO escape to the R5.017 mAb, we assessed the functional impact of the mutations herein reported on PfRH5 complexing with either binding partner PfCyRPA, receptor basigin or mAbs. Using a combination of *in-silico* prediction tools, we threaded these polymorphisms onto the structure of PfRH5 in complex with either PfCyRPA, BSG and some of the mAbs used in this study for which crystal structures have already been solved and reported^25,30,31^. Individual FASTA files with PfRH5 and individual SNP amino acid sequences were threaded through the crystal structure and the impact of the mutant versions of the protein was evaluated for predicted binding affinity of PfRH5 to its binding partner PfCyRPA, its erythrocyte receptor, Basigin, or the test human mAbs. The structure of PfRH5 used here comprises a truncated version of the protein in which the flexible N-terminus (residues E26 - Y139) and internal disordered loop (residues N248 - M296) regions were removed to enable crystallization^32^. Consequently, only seven out of the twelve SNPs reported here could be accurately mapped onto the PfRH5 structure by our model (**Fig. 6A**). Notably, these SNPs were evenly distributed between the N- and C-termini of the protein with C203Y, the most prevalent SNP reported here, occurring at the tip of the second helix around the BSG binding site, and F505Y present around the PfRH5-PfCyRPA interaction site (**Fig. 6A**). The difference in binding energy between the mutant and wild-type versions of PfRH5 was predicted by FoldX version 5.0 and was used to estimate the functional impact of each SNP on protein folding, or binding dynamics with PfCyRPA, Basigin, or mAbs (**Supplemental Table 2)**. Except for D249G, for which no structural information was available, all previously reported SNPs were predicted to have a mild impact on PfRH5 by either altering its conformation and stability (K221R, E306G, V371I and I407V) or enhancing its binding to BSG (**Fig. 6B-H**). On the other side, while no functional prediction was possible for majority of the novel SNPs (K51R, I60V, Y254C and F505Y), the impact of D243N, although near the interface of PfRH5-PfCyRPA, was not predicted to impact the structure.

**Figure 6:**
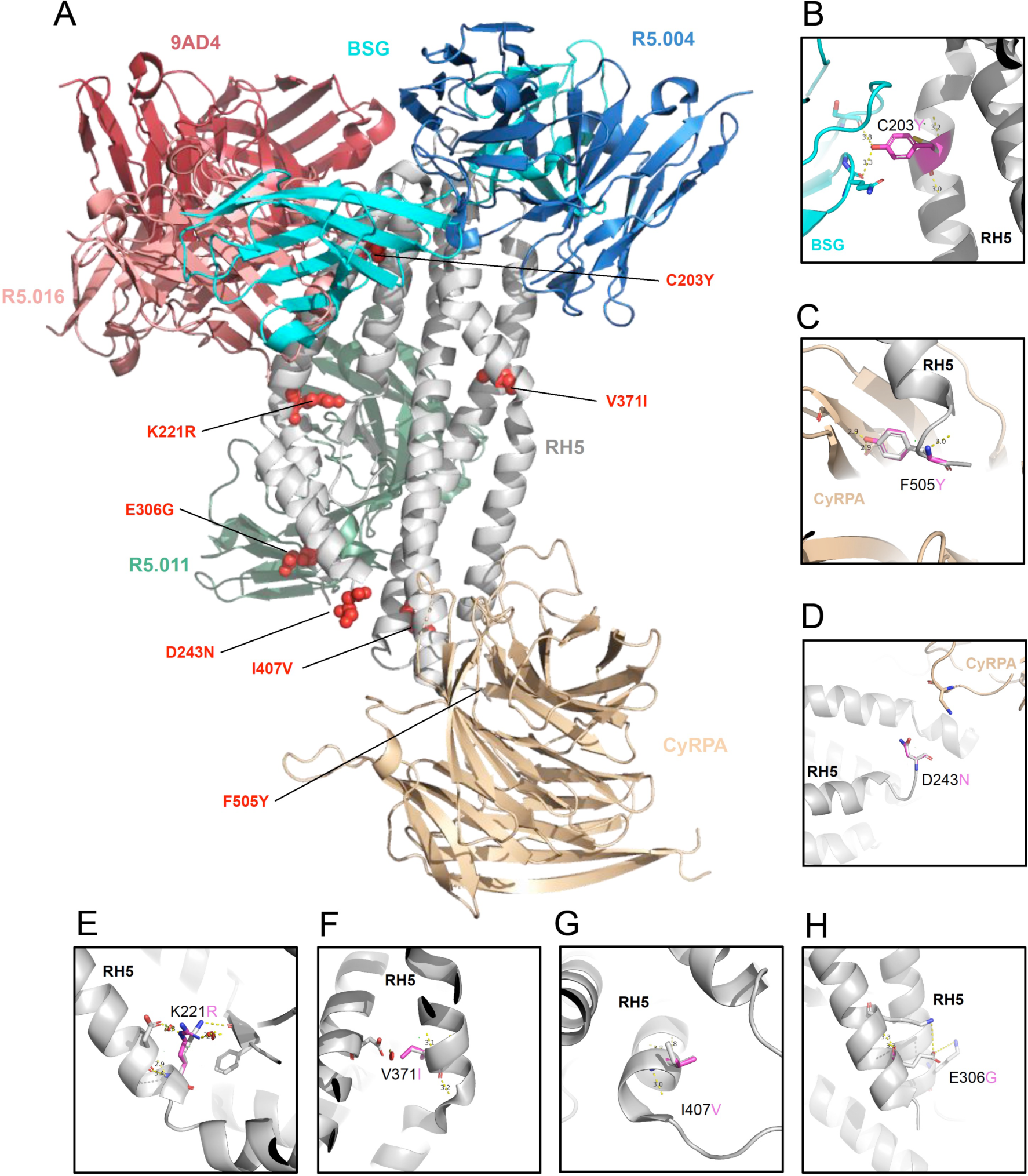
Structural threading of PfRH5-associated polymorphisms onto the BSG-PfRH5-PfCyRPA complex. (**A**) Distribution of identified SNPs, red spherical shapes, onto the PfRH5 structure. The model presented here was built by superposing the structure of PfRH5 in complex with BSG (4U0Q), PfCyRPA (6MPV), 9AD4 (4U0R), R5.004 and R5.016 (6RCU) and R5.011 and R5.016 (6RCV). BSG, PfRH5 and PfCyRPA ribbons are shown by light aqua, grey and light orange, respectively. Ribbons of the 9AD4, R5.004, R5.011 and R5.016 mAbs are depicted in dark red, blue, green and light red, respectively. (**B-H**) Sub panels highlighting the predicted effect of selected SNPs, the black dotted lines represent hydrogen bonds or salt bridges. From top-left to bottom-right: SNPs that potentially impact binding of PfRH5 and BSG (**B**), to enhance binding of PfRH5 to PfCyRPA (**C**), with no predicted function (**D**) or to alter the structure of PfRH5 (**E-G**).

## Discussion

PfRH5 is currently the lead blood-stage malaria vaccine candidate, with recent promising clinical trial outcomes^17,18,24,33^. Malaria vaccine development, however, has long been hampered by the extensive genetic diversity of naturally circulating parasite populations. Early testing of the functional impact of natural genetic diversity is critical to the development of a strain-transcendent PfRH5-based malaria vaccine. While vaccine-induced mAbs to PfRH5 are associated with varying levels of neutralizing activity against *P. falciparum* laboratory lines from different geographical locations^25^, these had not been tested on *P. falciparum* clinical isolates, the ultimate target of any given malaria vaccine. Importantly, we and others have recently reported an emergence of rare PfRH5 variants from naturally circulating *P. falciparum* clinical isolates across Africa^34–38^.

Having earlier reported the near-exclusive invasion inhibition in clinical isolates using low concentrations of antibodies to the PfRH5 receptor, Basigin^36^, here, we aimed to assess the effectiveness of a previously characterized panel of vaccine-induced anti-PfRH5 mAbs on *P. falciparum* clinical isolates. The mAbs used here had been classified into different groups based on their GIA profiles^25^, Our present findings complement these reports and constitute, to our knowledge, the first report on the susceptibility of *P. falciparum* clinical isolates from natural infections to vaccine-induced anti-PfRH5 mAbs. Using a flow cytometry-based *ex-vivo* GIA assay, we showed these mAbs have broadly neutralizing activity resulting in a dose-dependent inhibition of *P. falciparum* clinical isolates; and more interestingly, in addition to the earlier reported dose-dependent susceptibility of *P. falciparum* parasites to anti-PfRH5 mAbs^22,25^, our findings accurately reproduced the categorization of these mAbs into three main groups with distinct GIA profiles^25^. Interestingly, we also found a comparable level of susceptibility of the clinical isolates relative to the reference clone 3D7, which harbors the vaccine allele used for immunization, therefore consolidating earlier studies reporting the clear evidence of PfRH5-associated strain-transcending functional activity^14,20,23,25^. This outcome is particularly promising as, if confirmed, will help rule out the earlier observed and most feared strain-specific efficacy associated with previous vaccine candidates^39^.

Given the extremely rapid and efficient process of merozoite invasion of erythrocytes, any malaria vaccine targeting the blood-stage of the parasite’s life cycle needs to trigger higher titers of antibodies or a combination of antibodies that can synergize together to achieve protection even at low concentrations. While the former is difficult to achieve^23^, there is recent evidence of synergistic interactions between anti-PfRH5 antibodies leading to enhanced potency^25,40^. Consequently, we assessed the earlier reported synergistic profile of R5.011^25^ and found that this mAb was able to potentiate the neutralizing activities of the least potent mAbs. Interestingly, while the majority of these interactions were additive, we also noted some antagonistic interactions between R5.011 with either R5.001, R5.007 or R5.008. These contrasting results however involved only two clinical isolates, which showed antagonistic relationships across all combinations and warrant further investigation to better ascertain the contributing factors to the observed phenotypes. Encouragingly, while we observed some variation in the clinical isolates’ susceptibility to anti-PfRH5 mAbs, relative to our set threshold, none of these showed a complete loss of GIA susceptibility across all mAbs. On the other hand, the observed differences in percent GIA between clinical isolates and the 3D7 reference clone, although not significant, could possibly be attributed to donor-to-donor erythrocyte variance, a major contributor in the GIA error of the assay (EoA)^28^. Notably, with respect to our set range of starting parasitemia (0.5-1%), parasites were diluted with fresh erythrocytes from a single donor only in 13 out of the 22 assays performed, while the remaining assays were directly performed with patient-derived erythrocytes.

Given parasite genotype can impact susceptibility to vaccine-induced antibodies, we sought to investigate the breadth of PfRH5 genetic diversity in our study population. As in our previous study, we implemented a deep amplicon sequencing of the PfRH5 gene using a sensitive discovery threshold of 2% prevalence^35^. This analysis resulted in the discovery of 12 SNPs, the majority of which were present at low prevalence, and detected in single isolates. Interestingly, we observed a deficit of the wild-type allele (the vaccine allele), represented in only one-third of the isolates, while the most common mutant allele was represented by the C203Y mutation, therefore confirming previous findings^35,36,41,42^. Moreover, 5 out of these 12 SNPs were novel and, to our knowledge, have not been previously reported. Furthermore, as our genotyping experiment revealed an average of 3 parasite genotypes per isolate; we sought to assess the frequency of the identified SNPs within the complex mixture of parasite genomes per isolate using our previously described approach^35^. This analysis revealed that the majority of the SNPs reported here were present at low variant read frequencies (<5%) in polygenomic infections, except for C203Y, which most of the time occurred at high variant read frequencies (Mean = 75.11%, ranges 4.1 – 100%). Interestingly, out of the 70 SNPs identified in our previous study as novel^35^, only four (K51E, K221R, D249G and E306G) were found in this study. Notably, most of the novel SNPs identified in our studies occurred in regions of the protein reported to be internally disordered and not included in the crystal structure of PfRH5^32^. These regions include the N-terminal domain of the protein (residues E26 - Y139), which is cleaved through plasmepsin X activity^43^, and the internal disordered loop (residues N248 - M296), both of which appear to routinely induce non-neutralizing antibodies^22,25,26,44^. These observations warrant further investigation on the spatiotemporal mapping of PfRH5*-* associated polymorphisms and genetic relatedness of circulating parasite populations to ascertain the contribution of this diversity in the susceptibility of the parasites to vaccine-induced antibodies.

As recently reported by Farrell *et al.*^45^, PfRH5 does not change its conformation upon binding to PfCyRPA or BSG and, given the previous reports that neutralizing antibodies to PfRH5 primarily recognize conformational epitopes^25,40^, we sought to assess the impact of the identified SNPs on PfRH5 structure or its binding with PfCyRPA or BSG, as well as their functional impact on PfRH5 recognition by neutralizing antibodies. To this end, the identified SNPs were mapped onto the PfRH5 crystal structure in complex with binding partner PfCyRPA, receptor BSG or neutralizing mAbs for which crystal structures are available. Expectedly, no information was observed for K51E, K51R, I60V, D249G and Y254H, as the sequences mapping to these regions were truncated in the PfRH5 sequence used to generate the protein’s crystal structure. Nonetheless, our prediction data revealed that the remaining SNPs could affect PfRH5 in different ways, either by altering its structure, or enhancing its binding to CyRPA or BSG. Interestingly, no SNP was found to lie within known antibody-binding epitopes. We also noted the binding site of E306G to occur on the opposite face of PfRH5 to the R5.011 binding epitope, while the structural prediction showed no impact of this SNP on antibody binding. This observation, combined with that of isolate 400128 (in which this mutation is present) which along with 400113 showed moderately antagonistic interactions in combinatorial experiments, warrants further investigation. However, the functional impact of D243N mutation (present in 400113) was predicted as unknown by our model, hence the need for further functional assays (e.g. use of transgenic lines) to ascertain the contribution of this mutation to the reduced GIA susceptibility with 400413.

Although this study provides interesting insights on the efficacy of vaccine-induced mAbs to PfRH5 on *P. falciparum* clinical isolates, it also has some limitations that are important to note. First, the data reported here are gathered from a relatively small sample size from sites with similar transmission patterns and therefore might only cover some of the diversity with regards to areas with different endemicity. Moreover, we used a flow cytometry-based assay to collect phenotypic data using *ex-vivo* cultured isolates, which represent the closest parasite populations to that from natural infections. Although this strategy provides more reliable data with regards to the parasite *in vivo* phenotype relative to data obtained from culture-adapted parasites or laboratory lines, its limitation remains the fact that it is practically impossible to obtain data from distinct technical replicates of the assay, therefore, leaving room for other extrinsic sources of variation to affect assay outcome. One way to address this is to repeat the assays with culture adapted isolates, which still has limitations as it is known that only dominant clones of a given isolate will grow out and adapt^46^, therefore this does not guarantee data reproducibility. To mitigate such possible outcomes, and based on our previous data on potential sources of variation^47,48^, all assays were conducted using erythrocytes from a single unexposed donor for parasite dilution where needed, while all antibody dilutions and culture media were prepared fresh and used within 24 hours. Furthermore, as the goal was to test as many isolates as possible with a reasonable range of concentrations spanning previously defined EC_50_s for laboratory isolates, we were unable to test a wider range of mAb concentrations, and therefore unable to fully compare these data to those obtained with laboratory lines using the same mAb panel. Notwithstanding, the data described here revealed a limited impact of these challenges on the assay outcomes, as the bulk of the clinical isolates tested here presented relatively similar susceptibility profiles relative to the 3D7 reference line. Another limitation of this study worth highlighting is related to our sequencing strategy. Indeed, here we opted for deep targeted sequencing, which is a powerful tool to accurately identify rare variants occurring at very low frequencies within the parasite populations, however, due to its resulting short reads, we were unable to accurately assemble full-length PfRH5 gene haplotypes within a given polygenomic infection to compare their genetic relatedness to earlier reported parasite haplotypes^34–38^.

In summary, this study provides, to our knowledge, the first compelling evidence of the susceptibility of *P. falciparum* clinical isolates to vaccine-induced mAbs targeting the leading blood-stage malaria vaccine candidate antigen PfRH5, as well as the additive interaction between antibodies with different GIA profiles. Moreover, although our data seem to highlight the lack of functional impact of the identified diversity in the study population, the results from our *in-silico* prediction of the functional impact of these polymorphisms on the parasites’ susceptibility warrant further biochemical and functional genetic studies to ascertain their contribution on the overall parasite phenotype.

## Methods

### Sample collection and patient socio-demographic characteristics

This study was conducted in Kédougou, the Southeastern region of Senegal, where malaria transmission is highly endemic with a high transmission intensity associated with the rainy season from June to December. The study protocol was approved by National Ethics Committee of Senegal (CNERS) (SEN19/36), the regulatory board of the Senegalese Ministry of Health and the Institutional Review Board of the Yale School of Public Health (2000025417); and informed consent was obtained from all participants and/or their legal guardians. A total of twenty-two patients, aged 10 to 63 years, presenting with a positive *P. falciparum* RDT and microscope results were recruited from two healthcare centers, Bandafassi (N=7) and Dalaba (N=15), between July – August 2022. The participant eligibility to the study was assessed by the local healthcare practitioner and informed consent was obtained from patients who tested positive for *P. falciparum* single infection on a Pf/Pan (HRP/pLDH) Ag combo rapid diagnostic test (RDT) (Accessbio). A subsequent confirmation of *P. falciparum* mono-infection was performed using a Pf/VOM (HRP/pLDH) Ag combo rapid diagnostic test (RDT) (Accessbio). A venous blood draw (5ml) was performed in EDTA vacutainers (BD Bioscience) from all enrolled patients and transported to the laboratory for processing and diagnostic confirmation by microscopy. Infected erythrocyte samples with confirmed *P. falciparum* infection were separated from other blood components and used for parasite culturing, *ex-vivo* GIA assays and parasite DNA isolation.

### Monoclonal antibody production

The generation of the mAbs used here has previously been described^25^. Briefly, the variable heavy and light chains of the antibody-producing genes were amplified from single-cell-sorted plasmablasts isolated from peripheral blood mononuclear cells (PBMCs) of vaccinees from the first-in-human viral-vectored PfRH5 vaccine trial (NCT02181088)^17^. Cognate, paired heavy and light chains were cloned into separate human IgG vectors and transformed into *E. coli* Mix & go competent cells. The panel of mAbs used in this study included previously well-characterized human-vaccine-induced antibodies (R5.001, R5.004, R5.007, R5.008, R5.011, R5.016 and R5.017) as well as two chimeric mAbs (c2AC7 and c9AD4). These mAbs were expressed by transfection of Expi293F cells with paired plasmids encoding matched heavy and light chains using the ExpiFectamine 293 (Invitrogen) reagents and purified from culture supernatants using a protein G column. Purified antibodies were subsequently buffer-exchanged and resuspended at 1mg/ml in incomplete parasite medium (RPMI 1640 containing 25 mM HEPES, 0.1mg/ml Hypoxanthine and 50 μg/mL Gentamicin) and stored at −20°C until use.

### Parasite culturing and growth inhibition activity assays

*P. falciparum* clinical isolates were cultured at 4% hematocrit in complete parasite medium (RPMI 1640 containing 25 mM HEPES, 0.1mg/ml Hypoxanthine, 0.5% Albumax II, 2 mg/mL sodium bicarbonate and 50 μg/mL Gentamicin and 5% normal human serum). Human erythrocytes of blood group O^+^ from a single donor were used for all cultures and GIA assays. For *ex-vivo* GIA assays, parasites were seeded at 0.5-1% parasitemia at 2% hematocrit and co-incubated at ring stage with different concentrations of human mAbs against PfRH5 (150, 50, 25 and 10μg/ml), while a mouse IgG was used as an isotype control in all assays. Assays were set up in duplicates in a total volume of 40 µL in flat bottom 96 well half-well area plates with additional smear wells to monitor parasite growth; and incubated at 37° C in an atmosphere of 5% O_2_, 5% CO_2_ and balanced with N_2_. Cultures were harvested after a reinvasion rate of at least 95% was reached in the control wells. Parasite cultures were transferred into U-bottom 96 well plates and washed with 1X PBS-3% BSA (100μl/well) at 1500 rpm for 5 minutes. The red blood cell pellets were subsequently stained with 1/2000 Sybr Green in PBS for 20 minutes at room temperature in a shaking plate and washed twice with 1X PBS-3% BSA at 1500 rpm for 5 minutes. The pellets were finally resuspended into 200 µl 1X PBS and data were acquired using a Cytoflex cytometer (Beckman Coulter), where 100,000 events were recorded per well. Flow cytometry data outputs were analyzed to determine parasitemia in each well using the Flow Jo_v10.8.1 software. Finally, the FlowJo outputs were exported as Excel files and the percent GIA for each well was calculated from the invasion rate of each well relative to that of the IgG isotype control (Mouse IgG1 kappa mAb, abcam ab81032) at any given concentration. For each concentration, parasitemia from the duplicate test wells were averaged and invasion inhibition, relative to isotype control of the matched concentration, was calculated as follows: %GIA = 100-[(Average Percent invasion in anti-PfRH5 mAb wells)/ (Average parasitemia in IgG1 isotype control wells) * 100]. The graphs were plotted using the GraphPad Prism version 1.0.2 software.

### PCR amplification and next-generation analysis

DNA was extracted from ring-infected red blood cell pellets using the QIAmp DNA Blood Mini Kit (Qiagen) following the manufacturer’s instructions. The PfRH5 gene was amplified as previously described^36^ using the high-fidelity platinum taq (Invitrogen) and the amplicon size was resolved in a 1% agarose gel electrophoresis. Successfully amplified products were subsequently subjected to Next-generation Sequencing using the Illumina NovaSeq 6000 platform using the following procedure. Briefly, PCR amplicons were bead-purified and quantified by Qubit and Bioanalyzer before library preparation using unique dual indexes (UDIs), which associate each sample with a unique dual index for easy identification after sequencing. Indexed samples were subsequently bead-purified to select for that fragmented DNA of 200-300 bp sizes, which were then quantified with qPCR using Illumina’s KAPA Library Quantification Kit and normalized to a concentration of 4nM. These normalized samples were pooled into 8 sets of 12 samples each and bead-purified once more to further select DNA with the desired range of sizes, which were run on another KAPA qPCR to assess concentration. The concentration of these 8 sub-pools was normalized to 4nM and combined in equal parts to form one final pool, which was submitted to the Yale Center for Genome Analysis (YCGA) for High Throughput Next Generation Sequencing with a NovaSeq 6000 with targeted coverage of 500,000 reads per sample.

### Analysis of natural genetic diversity and structural threading

For each sample demultiplexed forward and reverse sequencing reads were obtained from the sequencing platform and imported into the Geneious prime software, where matched sequencing reads were paired using the Illumina, paired end-setting and subsequently trimmed using the BBDuk plug-in, with a minimum quality score of (Q) 30 and a minimum length of 75 base pairs set, as reads around 100 base pairs were expected. Trimmed sequences were mapped to the 3D7 reference PfRH5 sequence (PF3D7 0424100) already annotated with all known synonymous and non-synonymous mutations. Sequence mapping was set for two iterations and coverage criteria were set at 1000 reads. The criteria for single nucleotide polymorphism (SNP) calling was set to a minimum frequency of 0.02 (2%) and 1000 read coverage. At least 3 individuals performed sequence data and SNP analysis for each sample using defined guidelines to ensure data quality. The structure of PfRH5 in complex with BSG (4U0Q), CyRPA (6MPV), 9AD4 (4U0R), R5.004 and R5.016 (6RCU) and R5.011 and R5.016 (6RCV) have been solved and are publicly available in Protein Data Bank (PDB). Identified SNPs were threaded onto the structure of PfRH5 in complex with either Basigin, CyRPA or monoclonal antibodies. The BSG-PfRH5-PfCyRPA-mAbs complex was built by superposing the PfRH5 in the BSG-PfRH5, PfCyRPA-PfRH5 and PfRH5-mAb complexes. PyMOL version 2.3.2 was used to predict the effect and to plot the structural location of each SNP^49^. Missing residues in antibodies, Basigin, and CyRPA were modeled in Maestro. FoldX v5 was used to predict the effects of SNPs on the stability of PfRH5 and binding affinity of antibodies, Basigin, and CyRPA.

### Statistical analysis

All data presented here were analyzed using GraphPad Prism version 10.0.2 (GraphPad Software Inc., California, US). The Fisher Exact test was used to compute differences between categorical variables. Mean GIA differences between isolates and across concentrations were assessed using Two-way ANOVA and, where significant, the Dunnett’s multiple comparison test was used for pairwise comparisons. Differences in GIA activities of antibody combinations were tested using the Wilcoxon matched-pairs signed rank test, while the Spearman rank correlation was used to assess relationships between variables. All tests were performed two-tailed and only a *p-value* of at least 0.05 was considered significant.

## Data Availability

All data produced in the present study are available upon request to the authors. Sequencing Reads associated with this study have been deposited in the NCBI SRA with the BioProject Accession: PRJNA1101747. GIA data and associated genotype data have been deposited in the Dryad database and are publicly available at: https://doi.org/10.5061/dryad.8931zcrzv

## Acknowledgements

We would like to thank Adama Diallo and Gerald Keita from Bandafassi, Souleymane Ngom from Dalaba, and all the healthcare workers at these sites for their partnership with Institut Pasteur Dakar. We would also like to thank the people of Kédougou for their participation and invaluable contributions to this work. We would like to thank the Yale Center for Genome Analysis (YCGA) and acknowledge the support of the NIH HPC equipment grant (1S10OD030363-01A1).

## Funding Sources

This work was supported by G4 group funding (G45267, Malaria Experimental Genetic Approaches & Vaccines) from the Institut Pasteur de Paris and Agence Universitaire de la Francophonie (AUF). AKB is supported by the Fogarty International Center of the NIH (K01 TW010496), National Institute of Allergy and Infectious Diseases of the NIH (R01 AI168238), LGT is supported by an ARISE grant from the African Academy of Sciences (ARISE-PP-FA-056). Research reported in this publication was supported by the National Institute of General Medical Sciences of the National Institutes of Health under Award Number 1S10OD030363-01A1 to YCGA. This study was additionally supported by National Institute of Allergy and Infectious Diseases (NIAID) grant R61 AI176583-01 to ZS.

## Author contributions statement

Project conception, management and coordination: LGT, AKB. Data collection and experimentation: LGT, AB, RL, MNP, AC, FD, SDS, AT, BDS, AM, AKB. Data analysis: LGT, AB, MNP, YG, ZS, AB, AKB. Contribution on reagents, materials, and analysis tools: SJD, KMc, DP, SPD, AM, IVW, LS. Paper writing: LGT and AKB. All authors read and approved the manuscript.

## Potential conflicts of interest

KMc and SJD are inventors on patent applications relating to RH5 malaria vaccines and/or antibodies. All other authors have declared that no conflict of interest exists.

## Data accessibility

Sequencing Reads associated with this study have been deposited in the NCBI SRA with the BioProject Accession: PRJNA1101747. GIA data and associated genotype data have been deposited in the Dryad database and are publicly available at: https://doi.org/10.5061/dryad.8931zcrzv

## Notes

### Competing Interest Statement

The authors have declared no competing interest.

### Author Declarations

The study protocol was approved by National Ethics Committee of Senegal (CNERS) (SEN19/36), the regulatory board of the Senegalese Ministry of Health and the Institutional Review Board of the Yale School of Public Health (2000025417); and informed consent was obtained from all participants and/or their legal guardians.

