## Supplemental Figures and Tables for "Vaccine-induced human monoclonal antibodies to PfRH5 show broadly neutralizing activity against *P. falciparum* clinical isolates"

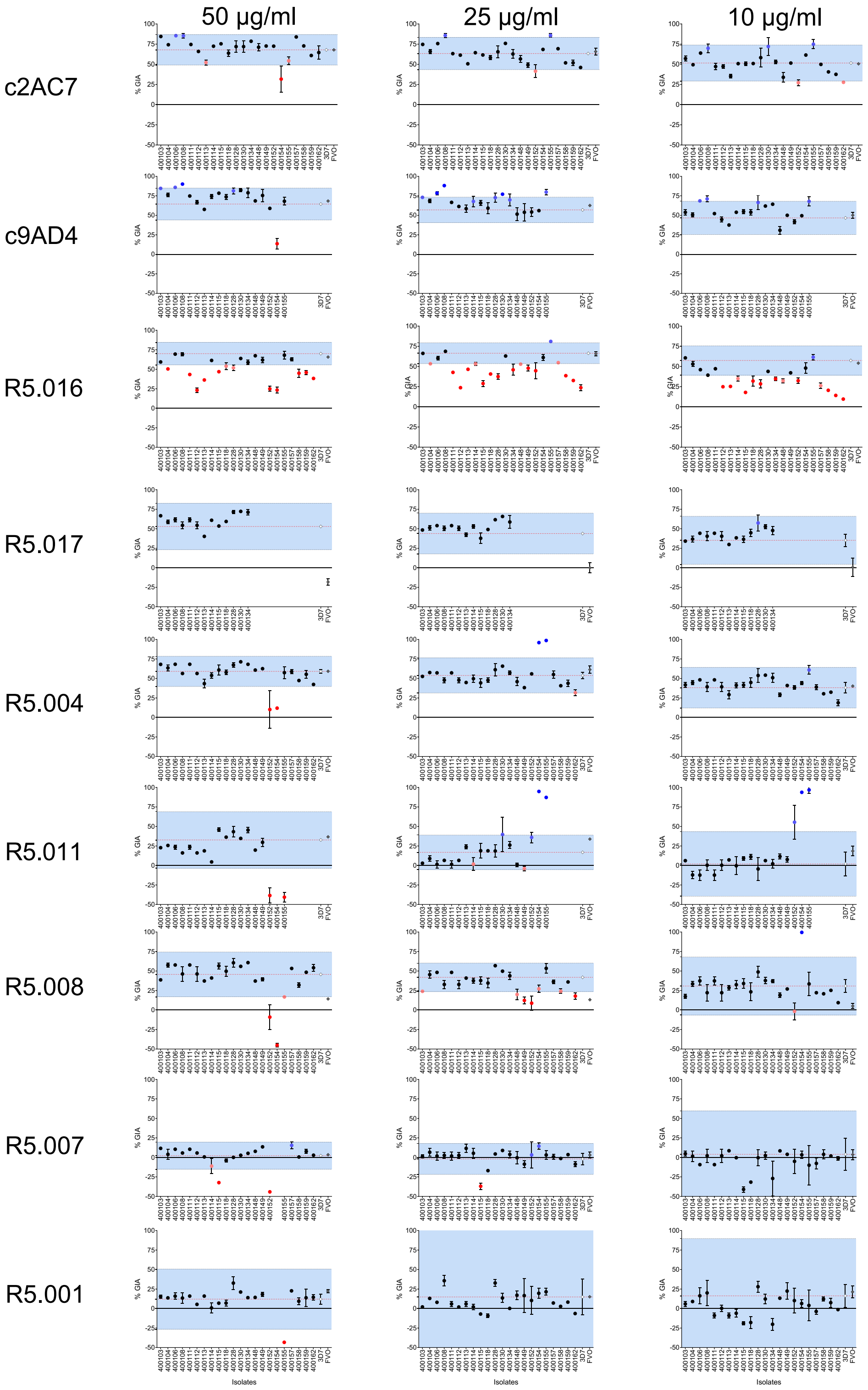


**Supplemental Figure 1: Variation of percent GIA ranges and immune susceptibility profiles of *P. falciparum* clinical isolates to PfRH5-vaccine-induced mAbs.** Dot plots of %GIA to anti-PfRH5 mAbs (Y-axis) of *P. falciparum* clinical isolates (X-axis) at different concentrations (50 µg/ml (**A**), 25 µg/ml (**B**) or 10 µg/ml (**C**)). The %GIA susceptibility ranges are defined as 3D7 mean %GIA (red dotted lines) ± 3SD 3D7 %GIA (black dotted lines). Black dots represent isolates lying within the defined susceptibility ranges. Red dots represent isolates reflecting a reduced GIA susceptibility phenotype (bright red) or laying at the borderline (dim red) of the lower limit of the defined susceptibility threshold. Blue dots represent isolates showing an increased GIA susceptibility (bright blue) or laying at the borderline (dim blue) of the upper limit of the defined susceptibility threshold.

| mAb | Concentration | Isolate | Genotype | p-value |
|---|---|---|---|---|
| **c2AC7** | 50 | 400106 | C203Y | 0.034 |
| | | 400162 | C203Y | 0.048 |
| **c9AD4** | 50 | 400128 | C203Y ; E306G | 0.046 |
| | 25 | 400108 | WT | 0.038 |
| | | 400130 | WT | 0.024 |
| | 10 | 400113 | D243N | 0.004 |
| **R5.016** | 50 | 400111 | K51E ; K51R ; C203Y | 0.011 |
| | | 400112 | K51E ; C203Y | 0.026 |
| | | 400113 | D243N | 0.047 |
| | | 400152 | C203Y | 0.039 |
| | | 400162 | C203Y | 0.012 |
| | 25 | 400112 | K51E ; C203Y | 0.005 |
| | | 400113 | D243N | 0.027 |
| | | 400118 | K51E ; C203Y | 0.015 |
| | | 400158 | C203Y ; V371I | 0.027 |
| | | 400159 | K51E ; K51R ; I60V | 0.017 |
| | 10 | 400106 | C203Y | 0.022 |
| | | 400108 | WT | 0.009 |
| | | 400112 | K51E ; C203Y | 0.017 |
| | | 400113 | D243N | 0.004 |
| | | 400115 | WT | 0.0007 |
| | | 400149 | C203Y ; I407V ; F505Y | 0.037 |
| | | 400158 | C203Y ; V371I | 0.002 |
| | | 400159 | K51E ; K51R ; I60V | 0.027 |
| | | 400162 | C203Y | 0.042 |
| **R5.004** | 50 | 400154 | C203Y | 0.013 |
| **R5.017** | 150 | 400113 | D243N | <0.0001 |
| | | 400130 | WT | 0.005 |
| | | FVO | S197Y | <0.0001 |
| | 50 | 400103 | D249G ; Y254C | 0.02 |
| | | 400113 | D243N | 0.03 |
| | | 400128 | C203Y ; E306G | 0.0006 |
| | | 400130 | WT | 0.0003 |
| | | 400134 | WT | 0.0008 |
| | | FVO | S197Y | <0.0001 |
| | 25 | 400128 | C203Y ; E306G | 0.001 |
| | | 400130 | WT | <0.0001 |
| | | 400134 | WT | 0.01 |
| | | FVO | S197Y | <0.0001 |
| | 10 | 400128 | C203Y ; E306G | <0.0001 |
| | | 400130 | WT | 0.001 |
| | | 400134 | WT | 0.042 |
| | | FVO | S197Y | <0.0001 |
| **R5.008** | 150 | 400114 | C203Y | 0.024 |
| | | 400155 | WT | 0.036 |
| | | 400158 | C203Y ; V371I | 0.024 |
| | | 400162 | C203Y | 0.012 |
| | 50 | 400154 | C203Y | 0.002 |
| | | FVO | S197Y | 0.025 |
| | 25 | 400128 | C203Y ; E306G | 0.038 |
| | | 400128 | C203Y ; E306G | 0.007 |
| **R5.011** | 50 | 400108 | WT | 0.049 |
| | | 400112 | K51E ; C203Y | 0.049 |
| | | 400154 | C203Y | 0.002 |
| | 25 | 400103 | D249G ; Y254C | 0.029 |
| | | 400154 | C203Y | 0.001 |
| | | 400155 | WT | 0.014 |
| **R5.007** | 50 | 400103 | D249G ; Y254C | 0.002 |
| | | 400115 | WT | 0.008 |
| | | 400152 | C203Y | 0.034 |
| **R5.001** | 150 | 400103 | D249G ; Y254C | 0.03 |
| | | 400108 | WT | 0.033 |
| | | 400155 | WT | 0.041 |
| | | FVO | S197Y | 0.046 |
| | 50 | 400154 | C203Y | 0.026 |
| | 10 | 400115 | WT | 0.024 |
| | | 400162 | C203Y | 0.033 |

**Table S1: Statistic table showing clinical isolates with a significant difference of %GIA relative to the 3D7 reference clone.** P-values were computed using the Dunnett's multiple comparison test following the Two-way ANOVA analysis and parasite genotypes were obtained using the Illumina NovaSeq 6000 next-generation sequencing platform.

| PDB ID | Complex | Mutation | ddG (kcal/mol) | Rh5 stability (kcal/mol) |
|---|---|---|---|---|
| **4U0Q** | RH5/Basigin | Y203C | 2.4296 | 0.753 |
| **4U0Q** | RH5/Basigin | E306G | 0 | 1.8518 |
| **4U0Q** | RH5/Basigin | F505Y | 0 | 0.1749 |
| **4U0Q** | RH5/Basigin | I407V | 0 | 1.1062 |
| **4U0Q** | RH5/Basigin | K221R | 0 | -0.2696 |
| **4U0Q** | RH5/Basigin | V371I | 0 | -0.124 |
| **4U0R** | RH5/9AD4 | Y203C | -0.0134 | 0.8367 |
| **4U0R** | RH5/9AD4 | E306G | 0 | 1.3719 |
| **4U0R** | RH5/9AD4 | F505Y | 0 | -0.0269 |
| **4U0R** | RH5/9AD4 | I407V | 0 | 1.3516 |
| **4U0R** | RH5/9AD4 | K221R | 0 | 0.6477 |
| **4U0R** | RH5/9AD4 | V371I | 0 | -0.5554 |
| **4U1G** | RH5/QA1 | Y203C | 0 | 0.868 |
| **4U1G** | RH5/QA1 | E306G | 0 | 1.2399 |
| **4U1G** | RH5/QA1 | F505Y | 0 | -0.018 |
| **4U1G** | RH5/QA1 | I407V | 0 | 0.7001 |
| **4U1G** | RH5/QA1 | K221R | 0 | 1.9648 |
| **4U1G** | RH5/QA1 | V371I | 0 | -0.646 |
| **6MPV** | Rh5/CyPRA/Ripr | C203Y | 0 | -0.298 |
| **6MPV** | Rh5/CyPRA/Ripr | E306G | 0 | 1.144 |
| **6MPV** | Rh5/CyPRA/Ripr | F505Y | 1.3743 | -0.424 |
| **6MPV** | Rh5/CyPRA/Ripr | I407V | 0 | 0.801 |
| **6MPV** | Rh5/CyPRA/Ripr | K221R | 0 | -0.384 |
| **6MPV** | Rh5/CyPRA/Ripr | V371I | 0 | 0.482 |
| **6RCU** | Rh5/R5.004 | C203Y | 0 | -0.84124 |
| **6RCU** | Rh5/R5.004 | E306G | 0 | 2.09274 |
| **6RCU** | Rh5/R5.004 | F505Y | 0 | 0.06114 |
| **6RCU** | Rh5/R5.004 | I407V | 0 | 0.89026 |
| **6RCU** | Rh5/R5.004 | K221R | 0 | 0.03218 |
| **6RCU** | Rh5/R5.004 | V371I | 0 | 0.7566 |
| **6RCV** | Rh5/R5.011 | C203Y | 0 | 1.811 |
| **6RCV** | Rh5/R5.011 | E306G | -0.0436 | 1.1167 |
| **6RCV** | Rh5/R5.011 | I407V | 0 | 0.7807 |
| **6RCV** | Rh5/R5.011 | K221R | -0.027 | -1.006 |
| **6RCV** | Rh5/R5.011 | V371I | 0 | 1.4678 |
| **7PHU** | Rh5/R5.015 | Y203C | 0 | 0.8163 |
| **7PHU** | Rh5/R5.015 | E306G | 0 | 2.763 |
| **7PHU** | Rh5/R5.015 | F505Y | 0.5901 | -0.5333 |
| **7PHU** | Rh5/R5.015 | I407V | 0 | 1.4872 |
| **7PHU** | Rh5/R5.015 | K221R | 0 | 0.2871 |
| **7PHU** | Rh5/R5.015 | V371I | 0 | 0.4805 |
| **6RCU** | Rh5/R5.016 | C203Y | -0.099 | -1.0154 |
| **6RCU** | Rh5/R5.016 | E306G | 0 | 1.7303 |
| **6RCU** | Rh5/R5.016 | F505Y | 0 | 0.0265 |
| **6RCU** | Rh5/R5.016 | I407V | 0 | 0.8954 |
| **6RCU** | Rh5/R5.016 | K221R | -0.0415 | 0.0724 |
| **6RCU** | Rh5/R5.016 | V371I | 0 | 0.7306 |

**Table S2. Predicted binding energy alternations for BSG and RH5 variant proteins:** Individual FASTA files with PfRH5 and individual SNP were threaded through the crystal structure and the impact of the mutant versions of the protein was evaluated for predicted binding affinity of PfRH5 to its binding partner PfCyRPA, its erythrocyte receptor, Basigin, or the test human mAbs. The structural effect of the PfRH5-associated SNPs was evaluated for predicted binding affinity between the mutant version of the RH5 protein and, binding partner PfCyRPA, the Basigin receptor or test mAbs. The binding energy alternation for SNPs and BSG were predicted by FoldX version 5.0. Predicted binding energies are shown for wild-type and mutant versions of the protein in Kcal/mol for each SNP. Changes between the two are shown as ddG (Kal/mol). A negative ddG indicates a predicted increase in binding and a positive ddG indicates a predicted decrease in binding.
